# No evidence for large causal effects of six air pollutants on non-suppurative otitis media: a two-sample Mendelian randomization study

**DOI:** 10.64898/2026.09.30.26364391

**Authors:** Ling Yang, Yang Liu

**Affiliations:** Department of Otolaryngology–Head and Neck Surgery, The First People’s Hospital of Fuzhou, Fuzhou 344000, Jiangxi, China

**Author notes:** Corresponding author: Yang Liu, Department of Otolaryngology–Head and Neck Surgery, The First People’s Hospital of Fuzhou, Fuzhou 344000, Jiangxi, China.

## Abstract

**Background:** Observational studies have linked air pollution to otitis media, but confounding and reverse causation preclude causal inference. We used two-sample Mendelian randomization (MR) to assess genetic causal effects of six air pollutants on non-suppurative otitis media, represented by otitis media with effusion (OME).

**Methods:** Exposure GWAS for PM2.5, PM2.5 absorbance, PM2.5-10, PM10, NO2 and NOx were obtained from UK Biobank (MRC-IEU), and the outcome was FinnGen R13 H8_NONSUPPNAS (12,397 cases, 464,237 controls). Inverse-variance weighting was primary, with MR-Egger, weighted median and mode, MR-PRESSO, leave-one-out and Steiger analyses, reverse MR and exploratory multivariable MR (MVMR). Asthma, allergic rhinitis and chronic rhinosinusitis served as positive controls, and three suppurative otitis media phenotypes served as negative controls.

**Results:** After false-discovery-rate correction, none of the six pollutants was significantly associated with OME; the sole nominal signal was PM10 (OR=1.555, 95% CI 0.947-2.553, P=0.081), which reversed with the 88-SNP pooled set (OR=0.64, 0.24-1.69, P=0.365). At 80% power, minimum detectable ORs ranged widely from 1.69 to 32.09; only PM2.5-10 and PM10 had values near 2, so moderate effects (OR 1.2-1.5) cannot be excluded. Confirmatory positive controls using phenotype-specific instruments were strongly positive, whereas negative-control outcomes showed no associations (all FDR>0.85). Every nominal reverse-MR signal had a significant MR-Egger intercept (the PM2.5 absorbance signal also showed significant heterogeneity and MR-PRESSO tests), indicating horizontal pleiotropy rather than reverse causation. MVMR was exploratory only, as conditional F statistics were below 10.

**Conclusions:** We found no evidence for large-effect genetic causal associations between the six air pollutants and OME. These null findings argue against large, lifetime-average causal effects, and the associations reported in observational studies may instead reflect residual confounding, shared comorbidity or differences in exposure windows.

## Introduction

Non-suppurative otitis media corresponds clinically to otitis media with effusion/middle-ear effusion (OME); the two overlap heavily in both clinical presentation and genetic research, and we use OME uniformly throughout this paper. The condition is characterised by non-suppurative middle-ear effusion and conductive hearing loss, is highly prevalent in children, and is one of the important treatable causes of conductive hearing loss and delayed speech and language development in children^[1]^; chronic recurrent disease can be complicated by adhesive otitis media, retraction pockets of the tympanic membrane and cholesteatoma^[1]^, and adds to the clinical and public-health burden^[2]^. Its aetiology involves multiple factors, including Eustachian tube dysfunction, infection, immune and inflammatory responses, allergy and anatomical abnormalities^[3]^. In recent years, ambient air pollution, a modifiable risk factor, has attracted increasing attention^[4]^, and available evidence suggests that it may be associated with the occurrence and recurrence of OME, although causality remains to be established.

Impaired Eustachian tube ventilation and drainage is the central process in the development of OME^[1]^. Air pollutants may induce inflammatory oedema of the nasopharyngeal mucosa and indirectly impair Eustachian tube function, leading to negative middle-ear pressure and effusion^[5][6]^. Previous epidemiological studies have mostly used observational designs with marked heterogeneity in conclusions: some cohorts reported associations of pollutant exposure with increased otitis media incidence^[4][7]^, whereas others reported only weak associations or associations confined to some pollutants^[8]^; such studies can rarely fully control for confounding by socioeconomic position, living environment, infection spectrum, allergy, smoking and season, and can seldom establish the temporal order of exposure and onset or exclude reverse causation. In addition, OME shows clear clinical comorbidity with asthma, allergic rhinitis and chronic rhinosinusitis^[9]^, and otitis media itself has substantial genetic susceptibility^[10]^, making it difficult for conventional regression to separate the independent effect of pollutants against a background of coexisting comorbidities.

Mendelian randomization (MR) uses genetic variants strongly associated with an exposure as instrumental variables; the random allocation of alleles at meiosis, which is independent of postnatal confounding, mitigates the confounding bias and reverse causation that affect conventional observational studies and provides a quasi-natural experimental framework for causal inference on environmental exposures^[11]^. Two-sample MR is conducted using publicly available genome-wide association study (GWAS) summary statistics and has been widely applied to explore the aetiology of complex diseases^[12]^. However, two-sample MR evidence on multiple air pollutants and OME remains limited: existing MR studies of otitis media have mainly focused on non-environmental exposures such as BMI^[13]^ and allergic rhinitis^[14]^, whereas MR studies of air pollution have mainly addressed inner-ear diseases^[15]^. In contrast, existing MR studies of air pollution have covered respiratory infections, including lower respiratory tract infections such as bacterial pneumonia and COVID-19^[16]^ and upper respiratory tract infections^[17]^, whereas middle-ear (OME) outcomes remain unexamined. Moreover, OME has substantial genetic susceptibility^[10]^, and environmental exposures and genetic factors may interact through pathways such as epithelial dysfunction, impaired mucociliary clearance, inflammation and oxidative stress, yet neither conventional observational studies nor existing MR studies can systematically integrate these dimensions. Studies that systematically integrate multiple pollutants while simultaneously evaluating reverse causation and the shared genetic architecture of comorbidities are still lacking^[18]^.

This study was reported in accordance with the STROBE-MR guideline^[19]^, using a two-sample MR design. We included GWAS of six air pollutants from the MRC-IEU OpenGWAS database^[20]^, generated from the UK Biobank cohort^[21]^, together with FinnGen R13^[22]^ non-suppurative otitis media case-control data. Inverse-variance weighted (IVW) estimation was used as the primary estimator, with the weighted median, MR-Egger, weighted mode, MR-PRESSO, Steiger directionality test and leave-one-out analysis for sensitivity analyses; reverse MR was used to assess reverse causation, and exploratory multivariable MR (MVMR) for PM10 was used to explore adjustment for confounding by the three comorbidities. Asthma, allergic rhinitis and chronic rhinosinusitis served as positive controls, and three suppurative otitis media phenotypes served as exploratory negative controls to examine phenotype specificity, aiming to clarify the aetiological relationship between air pollutants and non-suppurative otitis media from a genetic perspective and to provide evidence-based support for prevention and control.

## Materials and Methods

### Study design and data sources

We used a two-sample MR design: genetic instrumental variables were first selected from exposure GWAS data, and their effects on the outcome were then estimated in an independent outcome GWAS. All analyses used publicly available summary statistics, and no individual-level data were accessed; therefore, this secondary analysis required no ethical approval. The study was reported in accordance with the STROBE-MR guideline^[19]^. All GWAS participants were of European ancestry.

Instrumental variables for the six ambient air pollutants were obtained from the MRC-IEU OpenGWAS database^[20]^, which hosts GWAS summary statistics generated by the MRC-IEU pipeline from the UK Biobank cohort^[21]^. The six pollutants were: particulate matter with aerodynamic diameter ≤2.5 μm (PM2.5; GWAS ID ukb-b-10817, N = 423,796), PM2.5 absorbance (ukb-b-11312, N = 423,796), coarse particulate matter of 2.5–10 μm (PM2.5-10; ukb-b-12963, N = 423,796), particulate matter ≤10 μm (PM10; ukb-b-18469, N = 423,796), nitrogen dioxide (NO2; ukb-b-9942, N = 456,380) and nitrogen oxides (NOx; ukb-b-12417, N = 456,380). All pollutant phenotypes were measured at participants’ residential addresses using a Western European land-use regression model calibrated to 2010 monitoring data and integrating satellite remote sensing and chemical transport model information^[23][24]^; effect estimates in the source GWAS are expressed per one-standard-deviation (SD) increase in exposure. The continuous pollutant phenotypes had been inverse-rank normal transformed prior to the GWAS by the MRC-IEU pipeline.

The non-suppurative otitis media outcome was obtained from FinnGen R13 (phenotype H8_NONSUPPNAS; 12,397 cases and 464,237 controls), a nationwide Finnish biobank cohort with comprehensive registry-based phenotyping^[22]^. According to the Risteys phenotype definition (https://risteys.finngen.fi/endpoints/H8_NONSUPPNAS, accessed 19 September 2026), cases were defined by hospital discharge (HD) or cause-of-death (COD) records containing the whole ICD-10 chapter H65 (including H65.0–H65.4 and H65.9, covering acute/chronic serous, mucoid and other non-suppurative otitis media and unspecified non-suppurative otitis media) or ICD-9 code 381; no ICD-8 codes were available, and H68 (tubitis and obstruction of the Eustachian tube) was not included. Controls excluded all middle-ear and mastoid diseases (H8_MIDDLEMASTOID). The negative-control phenotype H8_MED_SUPP was based on the whole H66 chapter (including H66.9) and is mutually exclusive with the primary H65 outcome at the coding level; FinnGen allows the same individual to belong to different endpoints at different times, so a small number of individuals may overlap between the two case groups. The three comorbidities used in MVMR—asthma (J10_ASTHMA_EXMORE), allergic rhinitis (ALLERG_RHINITIS) and chronic rhinosinusitis (J10_CHRONSINUSITIS)—were also obtained from FinnGen R13. Negative-control outcomes comprised acute suppurative otitis media (H8_SUP_ACUTE, corresponding to H66.0; 14,387 cases; primary negative control), chronic suppurative otitis media (H8_CHRON_SUPOTITIS, corresponding to H66.1–3; 2,767 cases; secondary negative control), and the pooled phenotype suppurative and unspecified otitis media covering the whole H66 chapter including H66.9 (H8_MED_SUPP; 20,877 cases; used only for pooled sensitivity analyses).

Sample overlap and bias: the exposure GWAS for the primary analysis came from UK Biobank and the outcome GWAS from FinnGen R13; the two were independent with no substantial sample overlap, minimising weak-instrument and overlap bias arising from sample overlap in two-sample MR. Reverse MR (OME from FinnGen R13; pollutant outcomes from UK Biobank) likewise had no sample overlap. The three positive-control comorbidities and the OME outcome were all from FinnGen R13 and therefore overlapped, but their instruments were very strong (mean F ≈ 75–266), limiting overlap bias; the positive controls were consequently used only to verify the pipeline’s ability to detect true signals, and precise effect sizes were not interpreted. In MVMR, PM10 came from UK Biobank while the comorbidities and outcome came from FinnGen R13; the mixed sample structure and conditional F < 10 further restricted this analysis to an exploratory role.

### Instrument selection

Genetic instruments for each pollutant were selected through a two-stage procedure. First, single-nucleotide polymorphisms (SNPs) associated with each pollutant at the genome-wide significance threshold of P < 5×10⁻⁸ were identified. Because fewer than three independent SNPs were available for PM2.5-10 and PM10 at this threshold, the significance threshold for these two exposures was relaxed to P < 5×10⁻⁶. Second, linkage disequilibrium (LD) clumping was performed using the 1000 Genomes European reference panel with an r² threshold of 0.001 and a 10,000-kb window to retain mutually independent instrumental variables. Instrument strength was quantified by the F-statistic: the F-statistic for a single SNP approximates the square of its Wald test statistic for the exposure association, i.e., F ≈ (β/SE)², and overall instrument strength was reported as the mean F across all SNPs. The mean F was >20 for all exposures, indicating that weak-instrument bias was unlikely.

### Two-sample MR analysis

We considered the three core instrumental variable assumptions. First, relevance: the instruments were strongly associated with each pollutant, with mean and minimum F-statistics above 20 for all univariable analyses (Table 2). Second, independence: because alleles are randomly allocated at meiosis independently of postnatal environmental and socioeconomic circumstances, the instruments are expected to be independent of confounders of the pollutant–OME relationship; we assessed this indirectly through the pleiotropy, directionality and positive/negative-control analyses described below. Third, exclusion restriction: the instruments should affect OME only through the pollutant, which we evaluated using the MR-Egger intercept, MR-PRESSO, weighted median and weighted mode estimators, leave-one-out analyses and negative-control outcomes. These assumptions cannot be proven empirically and are addressed collectively by the sensitivity analyses.

The primary causal estimate used the inverse-variance weighted (IVW) method, which combines the per-SNP Wald ratio estimates by inverse-variance weighting; when Cochran’s Q indicated overdispersion, a multiplicative random-effects model was used (with standard errors scaled by Q/df), otherwise a fixed-effect model. Sensitivity analyses additionally reported the weighted median estimator (consistent even when up to 50% of instruments are invalid)^[25]^, MR-Egger regression, which detects and corrects directional horizontal pleiotropy through its intercept term^[26]^, and the weighted mode-based estimator based on the plurality assumption (which does not require valid instruments to form a majority). All analyses were performed with the TwoSampleMR package^[20]^: the package harmonises exposure and outcome data by aligning effect alleles, infers positive-strand orientation for palindromic SNPs from allele frequencies, and excludes ambiguous SNPs with frequencies near 0.5 that cannot be reliably oriented. No imputation was performed; SNPs that could not be harmonised were excluded.

### Sensitivity analyses

Horizontal pleiotropy was additionally assessed with the MR-Egger intercept test and the MR-PRESSO global test, the latter identifying and removing outlier instruments before re-estimating the causal effect^[27]^; the Steiger directionality test was used to verify that the assumed causal direction (pollutant → otitis media) was consistent with the variance explained by the instruments, and leave-one-out analyses were performed for each exposure to check whether a single SNP drove the results. Of note, MR-PRESSO could not be executed in the primary analysis pipeline owing to a function-parameter compatibility issue (the PRESSO fields in the summary results table are empty); the MR-PRESSO global tests in Table 4 were obtained independently with the MRPRESSO R package using exactly the same harmonised data (NbDistribution = 3000, set.seed(123)), and no outlier SNP was detected for any of the six exposures (zero outliers for all), so no outlier-corrected estimates were produced. Detailed scripts and parameters are provided in the code archive accompanying the manuscript and in the supplementary methods.

### Reverse MR and exploratory multivariable sensitivity analyses

Instrument strength in the multivariable setting was assessed with the conditional F-statistic^[28]^, and MVMR-IVW estimation followed the guidance of Sanderson et al.^[29]^. Given that the conditional F-statistics for PM10 in this analysis were all below 10 (see Section 3.6), MVMR was treated as an exploratory sensitivity analysis only and was not used for causal inference. The decision rule for reverse MR was prespecified: if the primary IVW analysis was non-significant, a positive Egger intercept would not be interpreted as evidence of reverse causation; if the primary IVW analysis was significant and the Egger intercept was also significant, directional pleiotropy would be indicated and the result interpreted with caution.

### Positive and negative controls

To assess the internal validity of the analysis pipeline, three positive controls were prespecified: asthma, allergic rhinitis and chronic rhinosinusitis—all well-recognised clinically related conditions of OME that should show strong causal signals if the data and methods are valid. Two sets of instruments were reported for the positive controls. The confirmatory analyses used only each comorbidity’s own independent genetic instruments extracted at P < 5×10⁻⁸ and clumped with a 10,000-kb window and r² = 0.001 (30 each for asthma and allergic rhinitis; 28 retained for chronic rhinosinusitis after harmonisation). The other set was the pooled candidate SNP set shared with MVMR (120 candidates; 115 after outcome harmonisation; 88 with complete data used in MVMR), which was not specific to any comorbidity and served only as a pipeline sanity check, not as confirmatory evidence. Negative-control outcomes (acute and chronic suppurative otitis media) were analysed with exactly the same six pollutant instruments as the primary analysis, to test whether the pollutant–OME associations were driven by non-specific pleiotropic effects of the instruments on middle-ear disease. Importantly, uniformly negative negative controls indicate only that no effect was detected at the available power: the lifetime-average effects captured by MR do not capture acute/developmental exposure windows. The null finding for acute suppurative otitis media (14,387 cases) indicates that no large lifetime-average effect was detected for this phenotype, whereas chronic suppurative otitis media had only 2,767 cases and limited power, and its results should not be over-interpreted. The negative controls were therefore exploratory phenotype-specificity analyses; they provide neither evidence excluding non-specific pleiotropy nor positive evidence for pathway specificity, and pipeline validity was judged solely from the positive controls.

### Statistical software

All analyses were performed in R (version 4.5.2), mainly using the TwoSampleMR package (version 0.7.9) and the MRPRESSO package (version 1.0). Multiple testing was corrected with the Benjamini–Hochberg false discovery rate (FDR), calculated separately within prespecified families of tests: the six pollutants in the primary analysis as one family, the six pollutants in reverse MR as one family, the six pollutants within each negative-control outcome as separate families, and each of the three positive controls based on its own independent instruments as a separate family. The study followed the STROBE-MR reporting guideline^[19]^. This study was not pre-registered; the analysis plan and scripts were finalised before the main analyses and archived at Zenodo (https://doi.org/10.5281/zenodo.22892731).

### Statistical power and minimum detectable effect

Given that null findings were the main results of this study, the statistical power and minimum detectable effect (MDE) for each exposure were evaluated using the instruments that actually entered the models. The proportion of exposure variance explained by the instruments was calculated as R² = Σ2EAF(1−EAF)β² (where EAF is the effect allele frequency and β is the SNP effect on the per-SD standardised exposure), and the IVW log-OR standard errors were taken from the empirical analyses. At two-sided α = 0.05 and 80% power, the log-OR corresponding to the MDE was (z₁₋α/₂ + z₁₋β)·SE; MDEs with Bonferroni correction for six tests (α = 0.05/6) were also reported, and power at a given assumed OR was calculated from the non-central χ² distribution. These estimates did not account for the additional uncertainty introduced by weak instruments, so actual power may be lower; per-instrument R², standard errors and MDEs are given in Table S8, with results summarised in Table 1.

**Table 1.** Statistical power and minimum detectable effects for MR analyses of six air pollutants and non-suppurative otitis media

| Exposure | No. of SNPs | R<sup>2</sup> (%) | IVW log-OR SE | MDE OR ($\alpha = 0.05$) | MDE OR (Bonferroni) | Power at OR = 1.3 (%) | Power at OR = 1.5 (%) | Power at OR = 2.0 (%) |
| --- | --- | --- | --- | --- | --- | --- | --- | --- |
| PM2.5 | 7 | 0.054 | 0.276 | 2.17 | 2.61 | 15.8 | 31.2 | 70.9 |
| PM2.5 absorbance | 4 | 0.030 | 1.238 | 32.09 | 74.31 | 5.5 | 6.2 | 8.7 |
| PM2.5-10 | 23 | 0.130 | 0.187 | 1.69 | 1.91 | 29.0 | 58.4 | 96.0 |
| PM10 | 27 | 0.150 | 0.253 | 2.03 | 2.41 | 17.9 | 36.1 | 78.2 |
| NO2 | 7 | 0.050 | 0.643 | 6.05 | 9.36 | 6.9 | 9.7 | 19.0 |
| NOx | 7 | 0.048 | 0.557 | 4.76 | 6.94 | 7.6 | 11.3 | 23.8 |

Note: MDE, minimum detectable effect (smallest OR detectable at 80% power); R<sup>2</sup>, proportion of variance in the per-SD standardised exposure explained by the instrumental variables; SE, empirical standard error of the IVW log-OR; the Bonferroni column applies correction for six tests ( $\alpha = 0.05/6$ ); power columns show statistical power (%) at the assumed OR.

## Results

### Instrumental variables

The numbers of independent instruments retained for the six pollutants were: 8 for PM2.5, 5 for PM2.5 absorbance, 24 for PM2.5-10, 30 for PM10, 8 for NO2 and 8 for NOx (Table 2). Mean F-statistics ranged from 23.06 to 37.83 and minimum F-statistics from 20.85 to 30.08, indicating that weak-instrument bias was unlikely in the univariable analyses. After harmonisation with the outcome data, 7, 4, 23, 27, 7 and 7 SNPs were retained for PM2.5, PM2.5 absorbance, PM2.5-10, PM10, NO_2_ and NOx, respectively. SNP loss during harmonisation did not exceed 30% for any exposure. Details of the instruments and F-statistics for each exposure and for reverse MR are given in Table S1; instrument information for the three covariates is given in Table S5; and the distribution of instrument F-statistics for each exposure is shown in Figure S6.

**Table 2.**
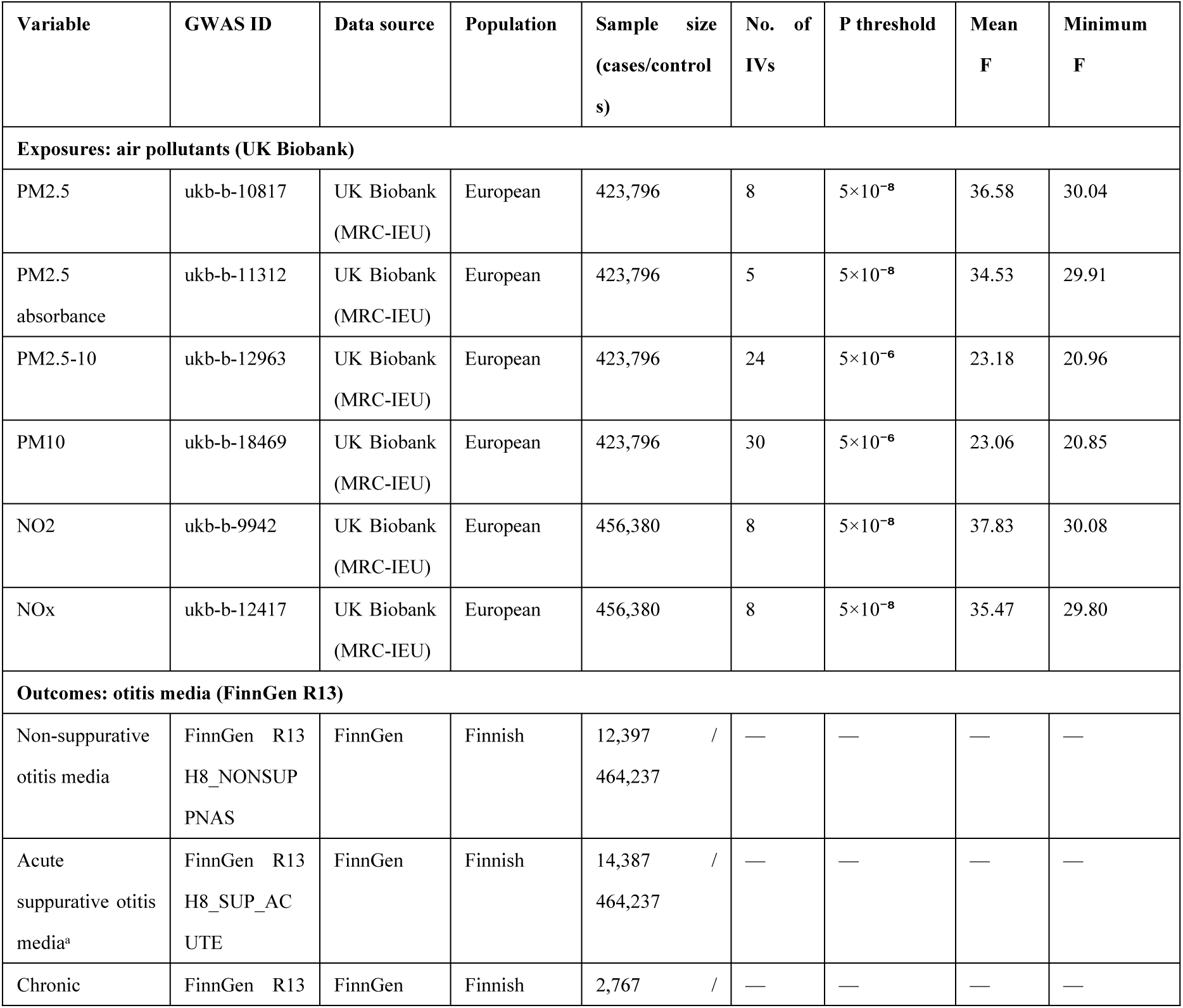
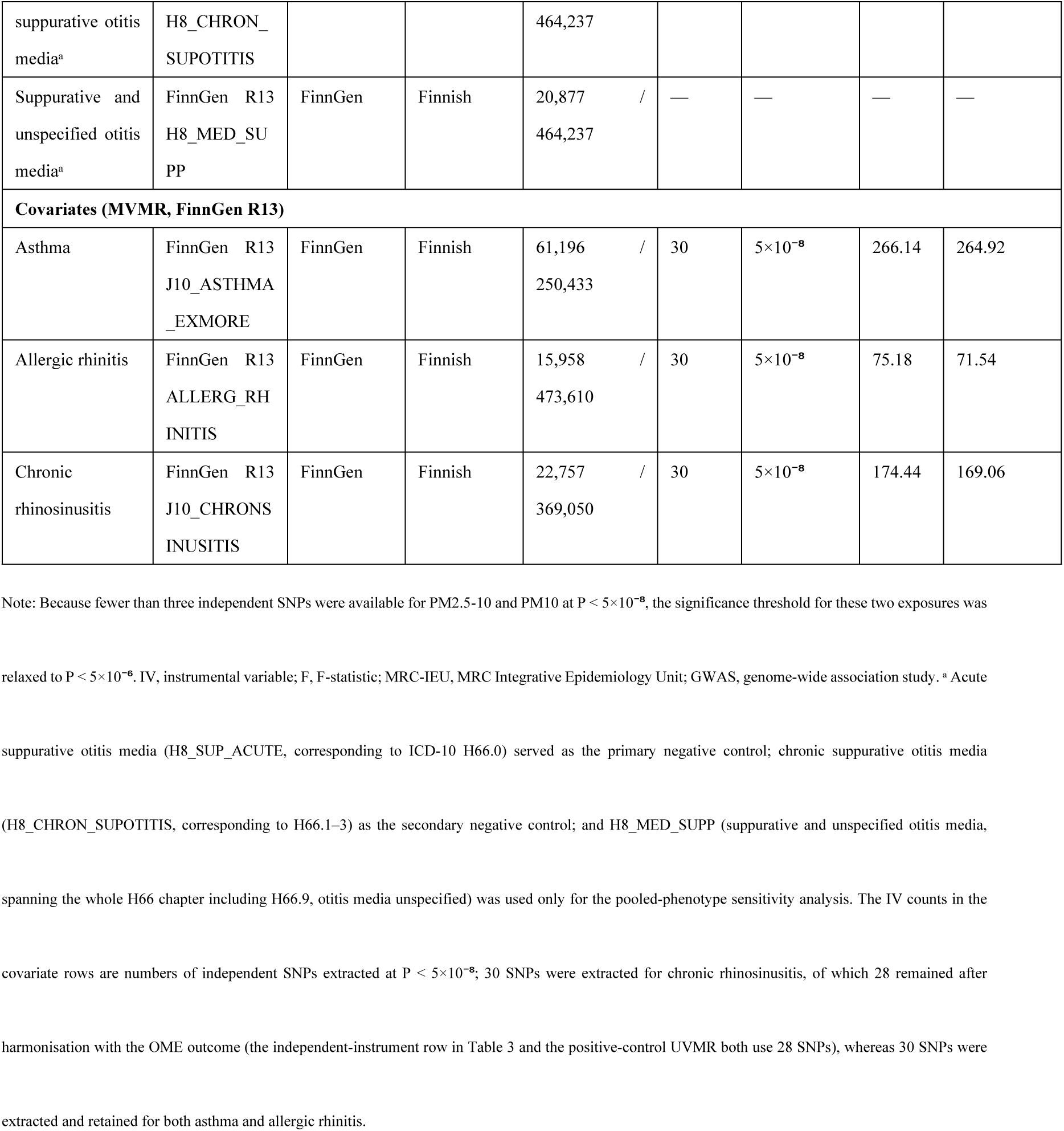
Characteristics of the GWAS summary datasets used in this study

### Primary MR analysis

IVW analyses showed no statistically significant associations between any of the six air pollutants and OME after FDR correction (Table 3, Figure 1). The ORs per SD increase were: PM2.5 1.021 (95% CI 0.595–1.755, P = 0.939), PM2.5 absorbance 0.914 (0.081–10.347, P = 0.942), PM2.5-10 1.097 (0.761–1.581, P = 0.620), PM10 1.555 (0.947–2.553, P = 0.081), NO2 2.499 (0.709–8.807, P = 0.154) and NOx 1.289 (0.433–3.838, P = 0.648). PM10 was the only pollutant with a nominally suggestive association, but after FDR correction P = 0.463 and it did not survive multiple-testing correction (Figure 1). Weighted median and weighted mode estimates were mostly directionally consistent with IVW and none reached statistical significance (Figure 2; complete results in Table S2); the methods gave opposite directions for PM2.5 absorbance, consistent with its only four instruments, significant heterogeneity and extremely wide confidence intervals (see Section 3.3). Of note, except for PM2.5-10, the other exposures could only detect effects of OR ≥ 2.0 at 80% power, and NO2, NOx and PM2.5 absorbance had less than 25% power even for OR = 2.0 (Table 1, Figure S7); the null findings should therefore be interpreted as a lack of evidence for large effects rather than exclusion of moderate effects.

**Fig 1.**
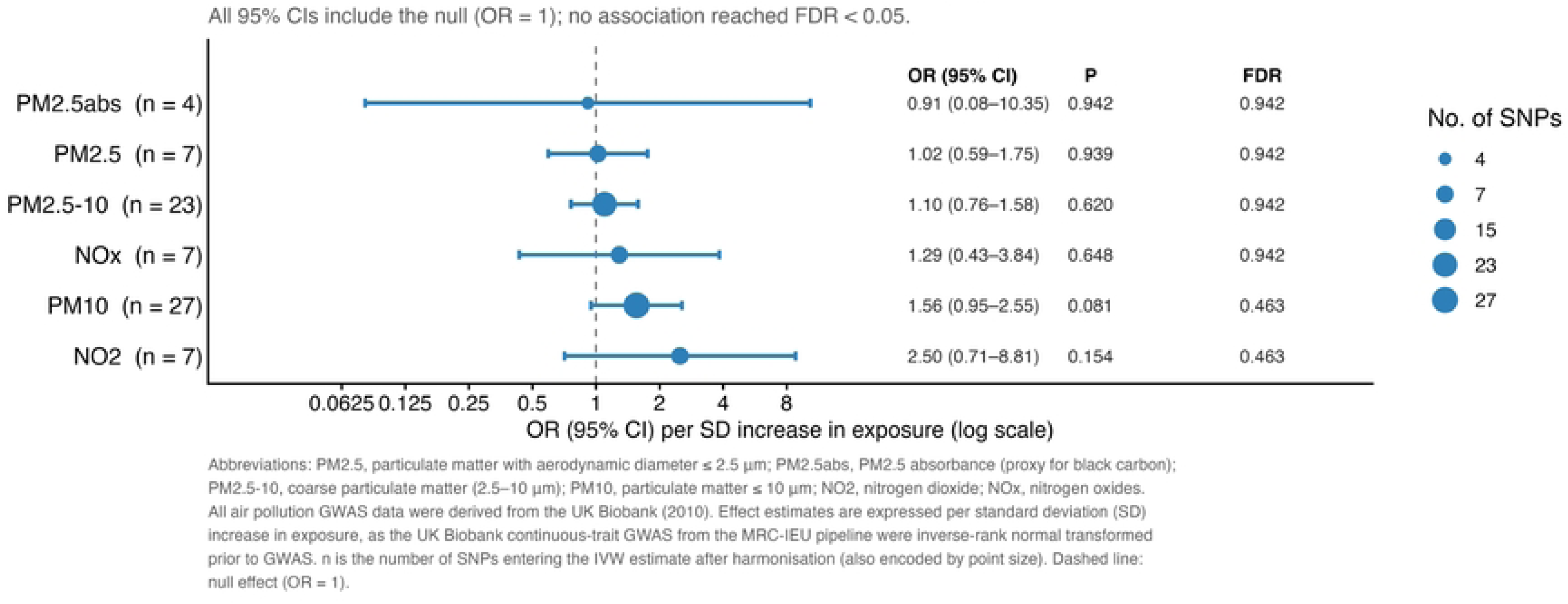
IVW MR estimates for six air pollutants on non-suppurative otitis media. Inverse-variance weighted odds ratios (ORs) with 95% confidence intervals (CIs), P values and false-discovery-rate (FDR) values are shown per standard-deviation (SD) increase in each pollutant. All 95% CIs include the null (OR = 1, dashed line) and no association reached FDR < 0.05. Point size encodes the number of SNPs entering the IVW estimate after harmonisation. PM2.5, particulate matter with aerodynamic diameter ≤2.5 µm; PM2.5abs, PM2.5 absorbance (proxy for black carbon); PM2.5–10, coarse particulate matter (2.5–10 µm); PM10, particulate matter ≤10 µm; NO2, nitrogen dioxide; NOx, nitrogen oxides. Exposure GWAS were UK Biobank continuous-trait GWAS from the MRC-IEU pipeline.

**Fig 2.**
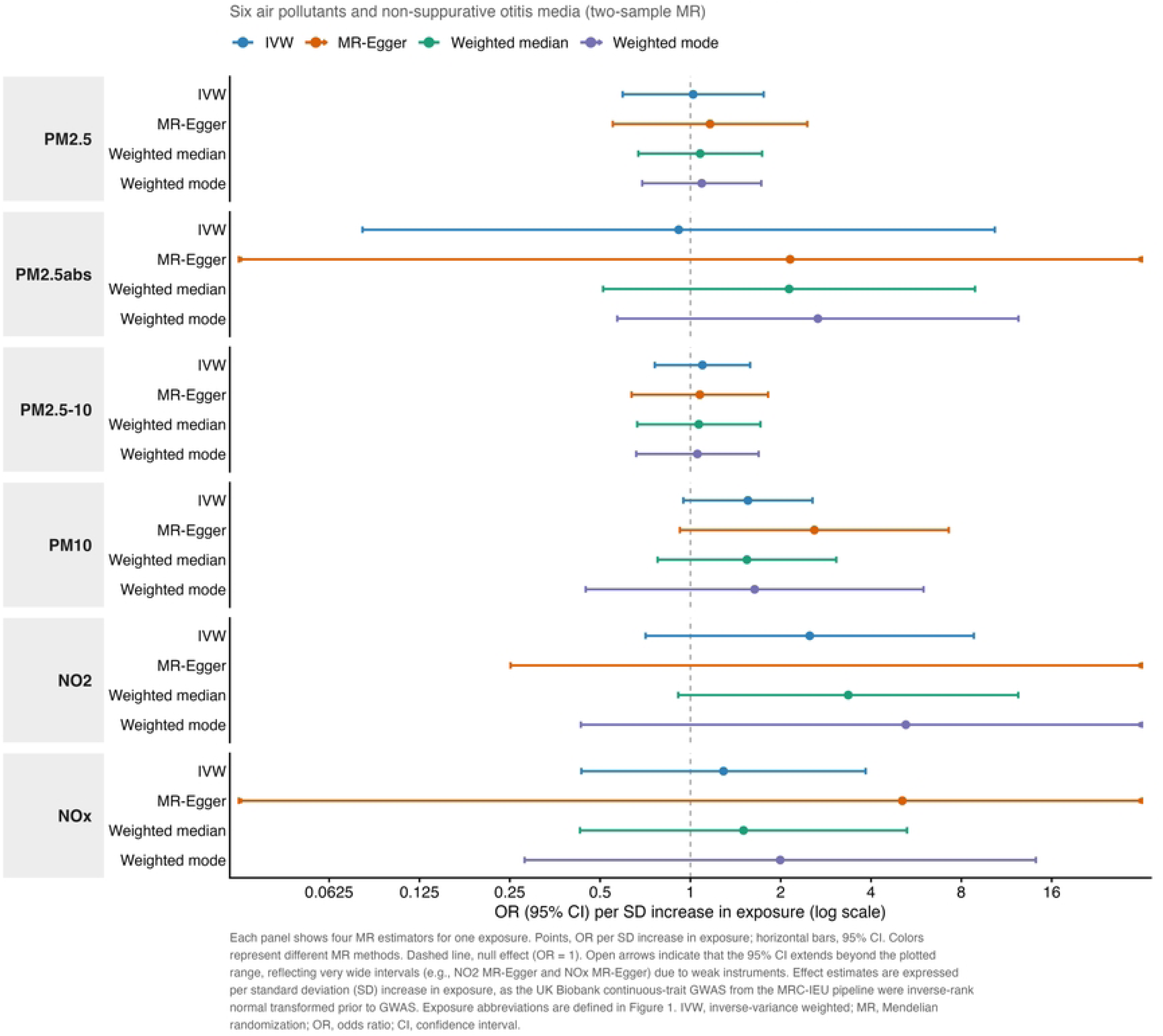
Consistency of causal estimates across four MR methods. ORs (95% CIs) per SD increase are shown for each pollutant from inverse-variance weighted (IVW), MR- Egger, weighted median and weighted mode estimators. The dashed line marks the null (OR = 1); arrows extending beyond the plotted range reflect very wide intervals from weak instruments (e.g. NO2 and NOx MR-Egger). Exposure abbreviations are defined in Fig 1.

**Table 3.**
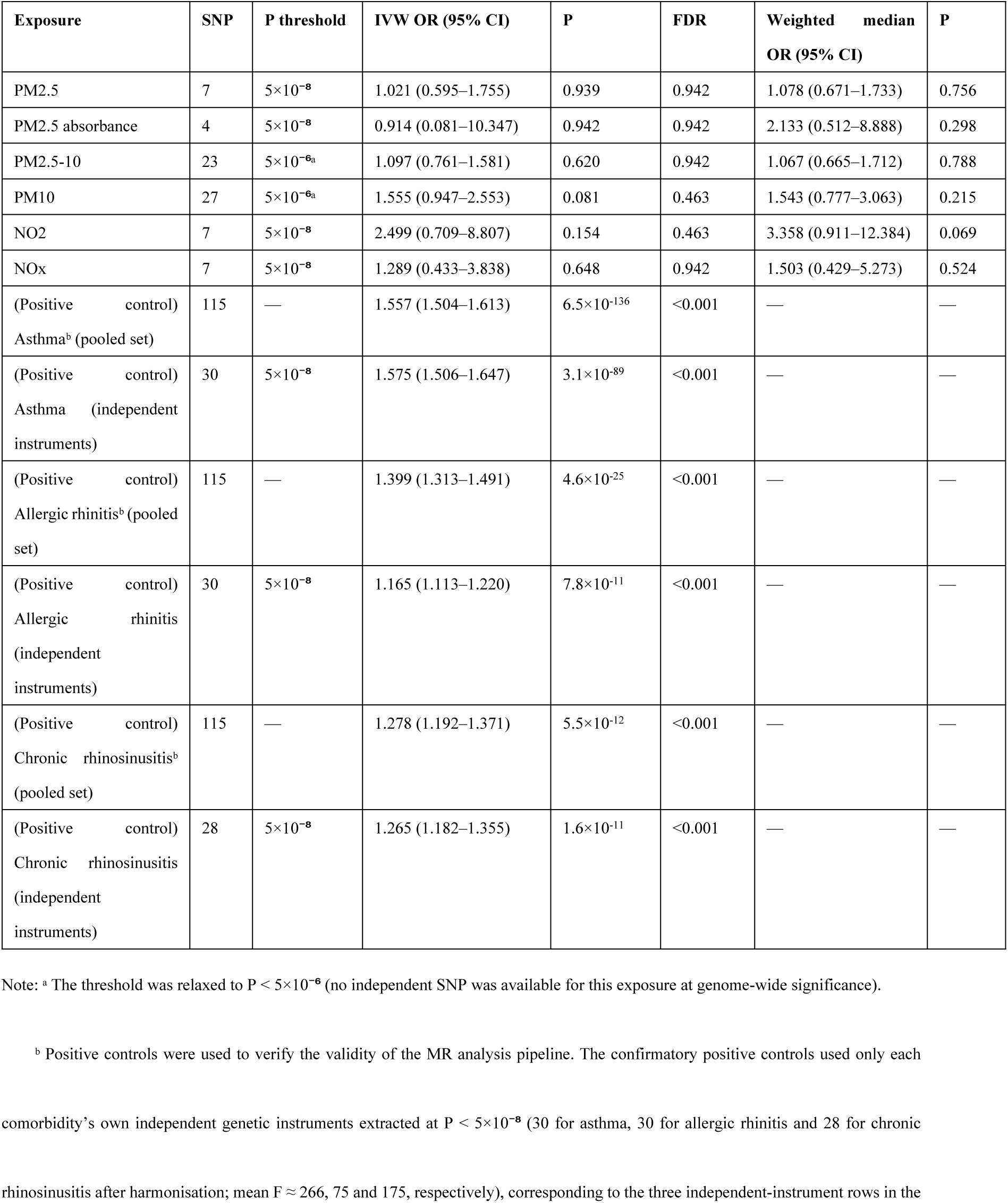
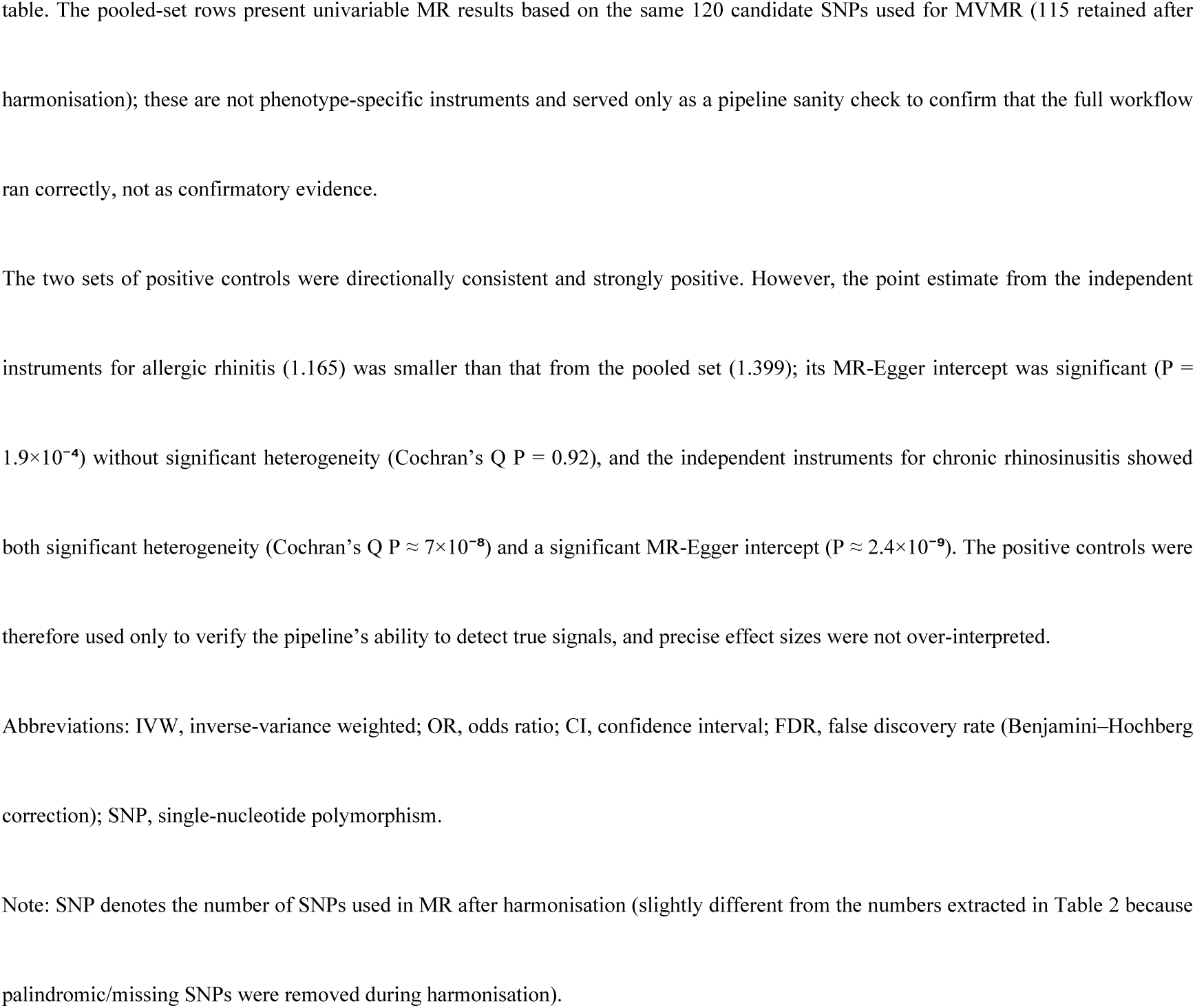
MR estimates for the associations of six air pollutants with non-suppurative otitis media

### Sensitivity analyses

Cochran’s Q tests showed significant heterogeneity only for PM2.5 absorbance (IVW Q P = 6.79×10⁻⁴; Table 4); its random-effects IVW standard error was scaled by Q/df = 5.70, and the wide confidence interval reflects between-instrument heterogeneity. Neither the MR-Egger intercept tests (all P > 0.23) nor the MR-PRESSO global tests reached significance, and no outliers were reported (Table 4); however, with few instruments for PM2.5 absorbance (4 SNPs) and NO2 (7 SNPs), these tests had limited power, and their null results should not be interpreted as evidence that pleiotropy is absent. The Steiger directionality test showed that the six exposures’ instruments explained more variance on the exposure side than on the outcome side, with all directions correct (P = 3.95×10^-8^ to 4.53×10^-50^, Figure 3D), supporting the assumed pollutant → OME direction. Leave-one-out analyses showed that no exposure’s estimate was driven by a single SNP; only PM2.5 absorbance fluctuated markedly after removal of rs59727727, consistent with its significant heterogeneity (Table S3). Single-SNP forest plots, funnel plots and scatter plots are provided in Figures S3–S4.

**Fig 3.**
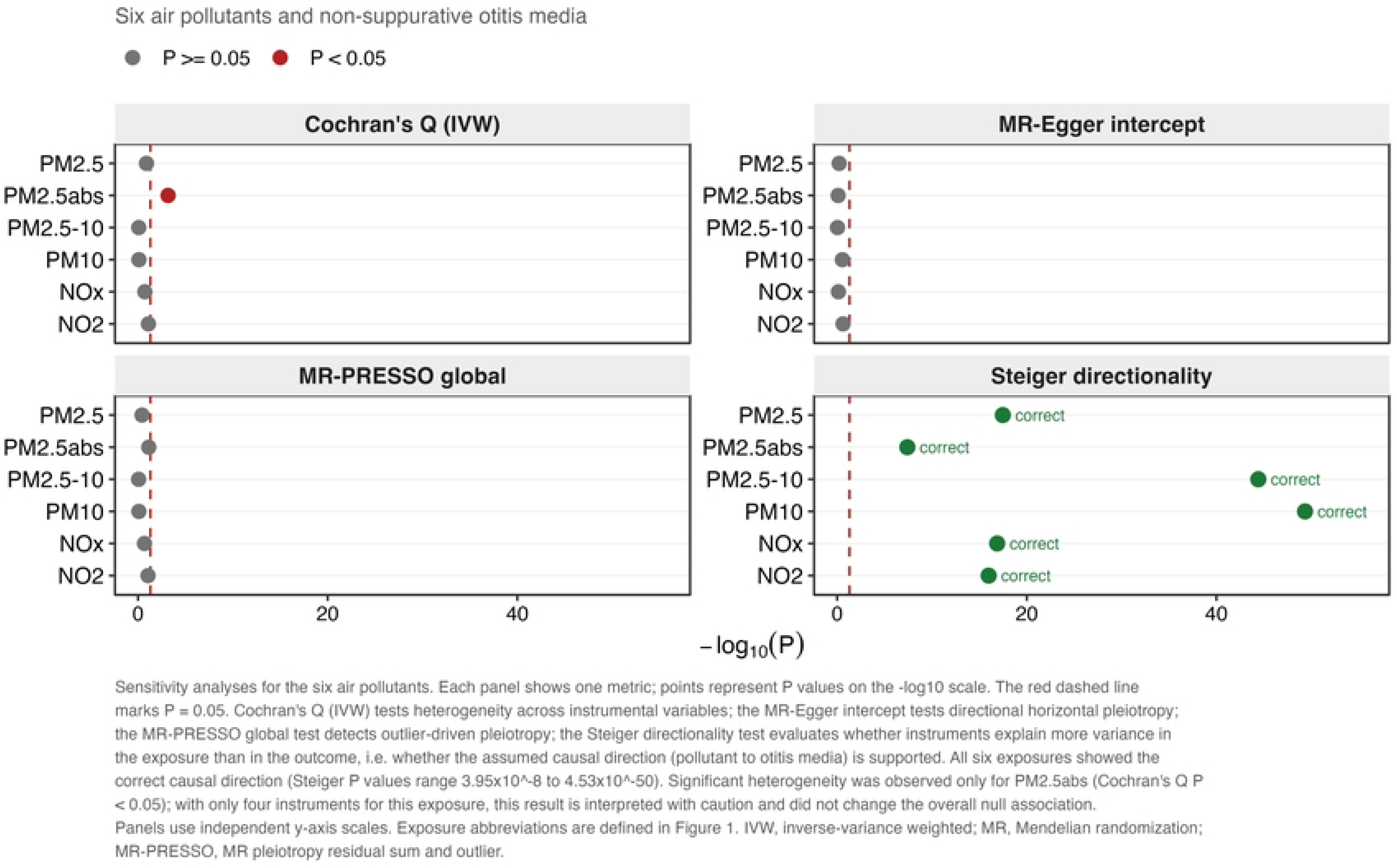
Sensitivity analyses: heterogeneity, pleiotropy and causal direction. The four panels summarise Cochran’s Q (IVW) for heterogeneity, the MR-Egger intercept for directional horizontal pleiotropy, the MR-PRESSO global test for outlier-driven pleiotropy and Steiger directionality for causal orientation. Points show P values on a −log10 scale; the red dashed line marks P = 0.05. Significant heterogeneity was observed only for PM2.5 absorbance (which had only four instruments) and did not alter the overall null association; all six exposures showed the correct causal direction (pollutant to otitis media).

**Table 4.**
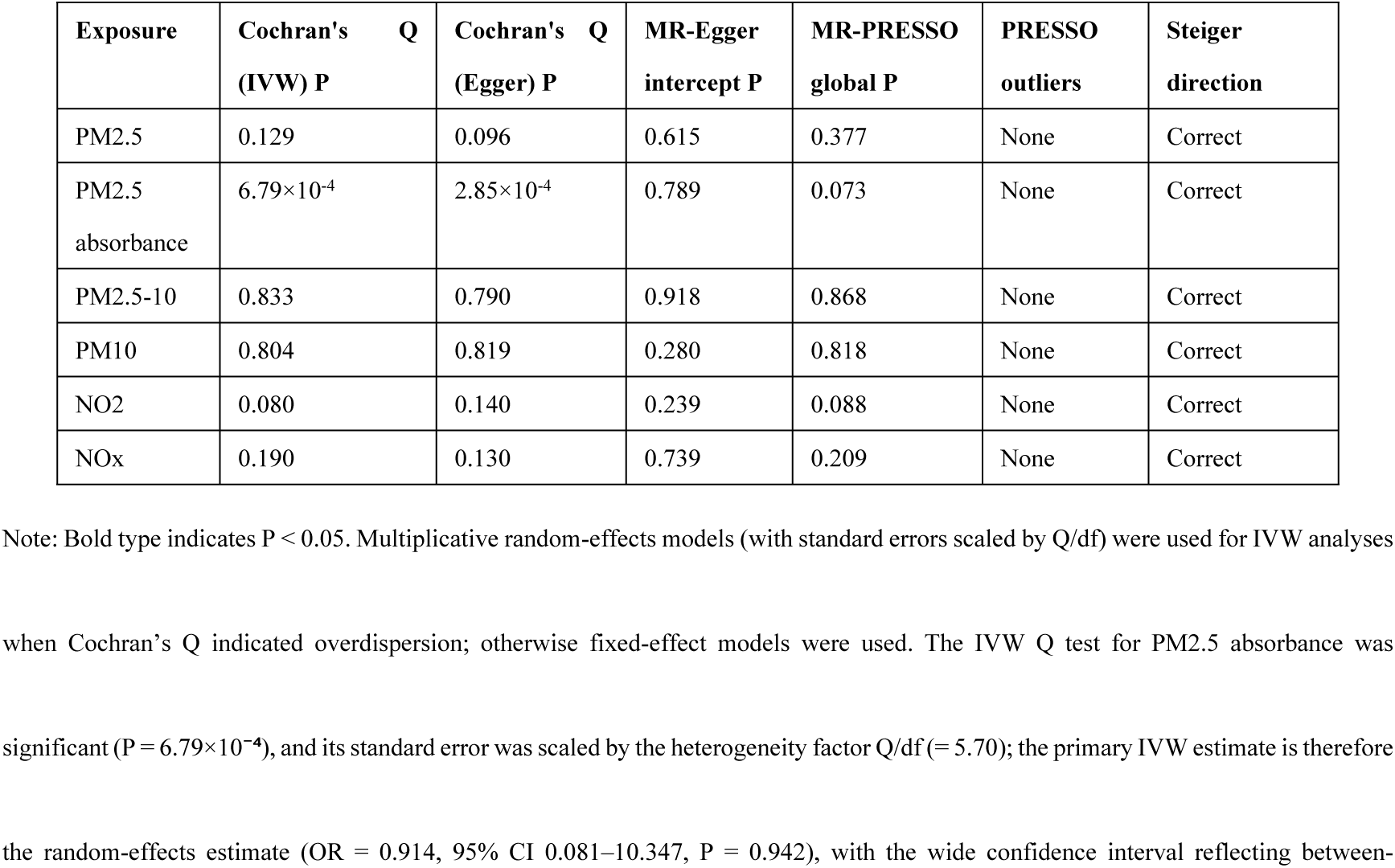
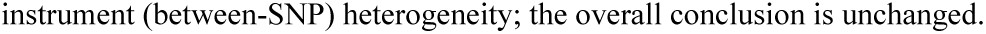
Sensitivity analyses for the associations of six air pollutants with non-suppurative otitis media

### Positive and negative controls

The confirmatory positive controls based on each comorbidity’s own independent instruments were all strongly associated with OME (all P < 0.001 after FDR correction; Table 3, Figure S1): asthma OR = 1.575 (95% CI 1.506–1.647), allergic rhinitis OR = 1.165 (1.113–1.220) and chronic rhinosinusitis OR = 1.265 (1.182–1.355), confirming that the outcome harmonisation and MR estimation pipeline could detect genuine causal associations. The 115-SNP pooled set shared with MVMR likewise gave positive results (asthma 1.557, allergic rhinitis 1.399, chronic rhinosinusitis 1.278), but this set was not specific to any comorbidity and was used only to confirm end-to-end pipeline connectivity, not as confirmatory evidence. The independent-instrument point estimate for allergic rhinitis (1.165) was smaller than that from the pooled set (1.399), with a significant MR-Egger intercept (P = 1.9×10⁻⁴) but no heterogeneity (Q P = 0.92); the independent instruments for chronic rhinosinusitis showed both significant heterogeneity (Q P ≈ 7×10⁻⁸) and a significant intercept (P ≈ 2.4×10⁻⁹). The positive controls were therefore used only to verify the pipeline’s detection ability, and precise effect sizes were not over-interpreted.

For the negative-control outcomes, IVW analyses of the six pollutants on acute suppurative otitis media (H8_SUP_ACUTE, primary negative control), chronic suppurative otitis media (H8_CHRON_SUPOTITIS, secondary negative control) and the pooled phenotype H8_MED_SUPP showed no statistically significant associations (Table S7, Figure S2): the IVW OR ranges for the three outcomes were 0.675–1.155 (smallest P = 0.415, FDR = 0.938), 0.443–1.162 (smallest P = 0.391, FDR = 0.956) and 0.527–1.146 (smallest P = 0.320, FDR = 0.859), respectively. Cochran’s Q, MR-Egger intercept and MR-PRESSO tests for each outcome were mostly P > 0.05, with no outlier instruments, and Steiger directions all supported pollutant → otitis media. The only borderline value was the Egger Q P = 0.044 for PM2.5 absorbance on H8_MED_SUPP, but its IVW Q P = 0.055 and primary IVW effect P = 0.320 did not change the null classification. Uniformly negative negative controls indicate only that no effect was detected at the available power; these were exploratory phenotype-specificity analyses and do not constitute evidence excluding non-specific pleiotropy. Their power limitations and role are described in Section 2.6, and pipeline validity remains based on the positive controls.

### Reverse MR

Reverse MR was performed using the 23 OME instruments that entered the final harmonised models (pollutants were per-SD standardised continuous phenotypes, so results are expressed as β rather than OR; instruments were clumped at P < 5×10⁻⁸ with a 10,000-kb window and r² = 0.001, yielding 30 independent candidate SNPs, of which 27 were present in the outcome GWAS and 23 entered the models after harmonisation; the 30 candidate SNPs had mean F = 59.86 and minimum F = 54.97). Genetically predicted OME had no significant effect on PM2.5 (β = 0.006, P = 0.226) or PM2.5-10 (β = −0.003, P = 0.581) (Table 5, Figure 4); nominally significant negative associations were seen for PM2.5 absorbance (β = −0.033, P = 1.58×10⁻⁵), PM10 (β = −0.012, P = 0.009) and NO2 (β = −0.013, P = 0.004), and NOx was a borderline signal (β = −0.008, P = 0.058). However, these reverse associations were all accompanied by significant MR-Egger intercepts (P = 1.5×10⁻⁴, 0.004, 0.007 and 0.009, respectively); the PM2.5 intercept was also nominally significant (P = 0.027), but its primary IVW effect was null (P = 0.226), which—following the prespecified rule in footnote ᵃ of Table 5—does not change the null conclusion. PM2.5 absorbance additionally showed significant heterogeneity (Q P = 2.14×10⁻⁵) and a significant MR-PRESSO global test (P < 3.33×10⁻⁴). These signals suggest that the reverse associations more likely reflect horizontal pleiotropy and shared confounding than genuine reverse causation. The Steiger test supported the reverse outcome → exposure specification for all six exposures (all directions correct; P = 1.26×10^-71^ to 1.01×10^-122^). Complete results are given in Table S6.

**Fig 4.**
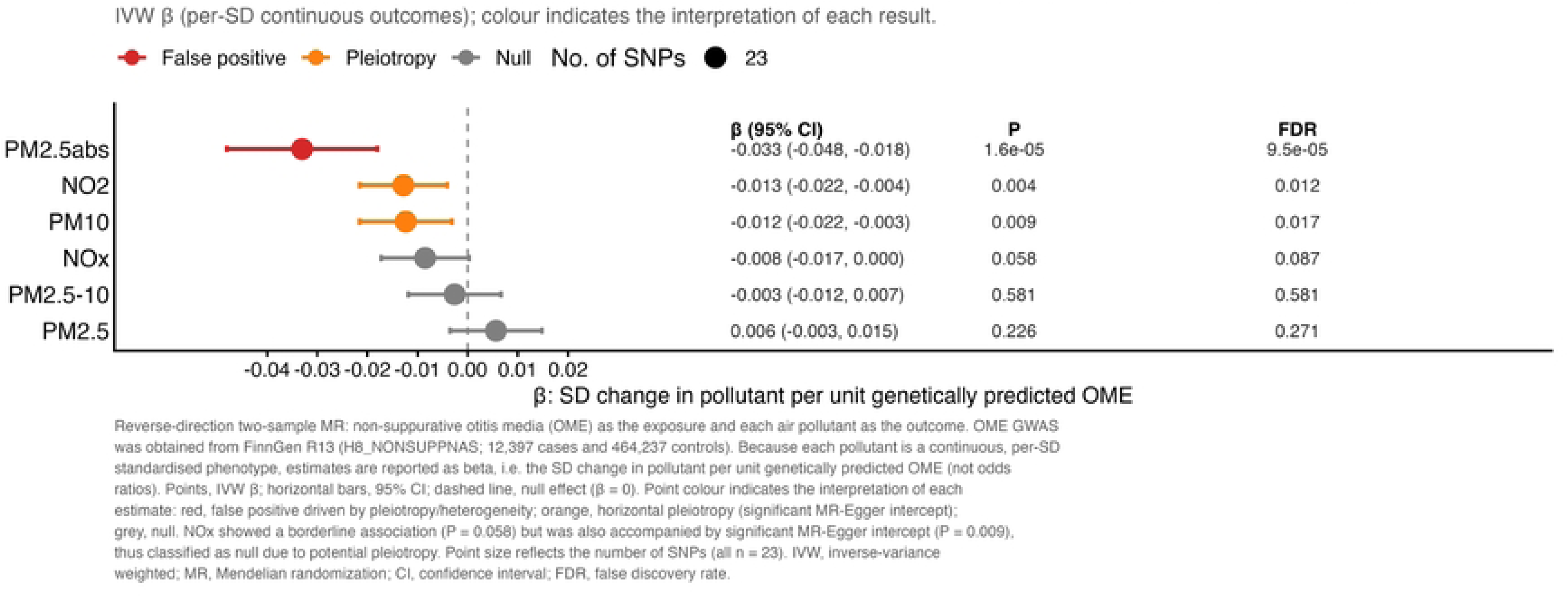
Reverse-direction MR: genetic liability to non-suppurative otitis media and six air pollutants. Inverse-variance weighted beta coefficients (SD change in each continuous pollutant per unit genetically predicted OME; 23 OME instruments) are shown with 95% CIs, P and FDR. Colour codes the interpretation: red, false positive driven by pleiotropy/heterogeneity (PM2.5 absorbance); orange, horizontal pleiotropy with a significant MR-Egger intercept (NO2, PM10); grey, null. The borderline NOx signal (P = 0.058) was likewise accompanied by a significant MR-Egger intercept and is treated as null.

**Table 5.**
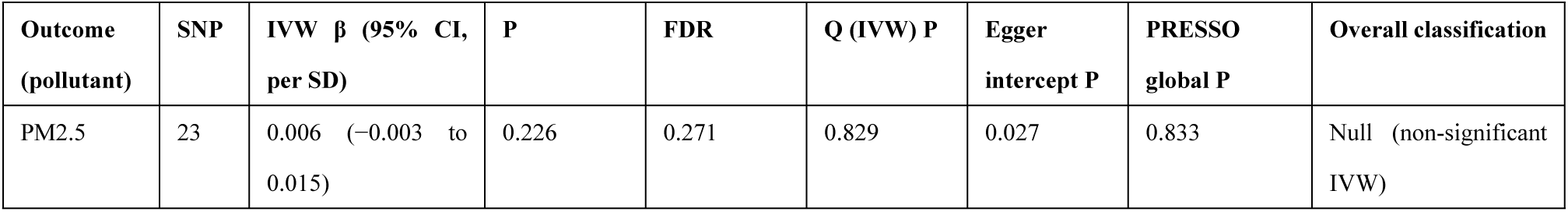
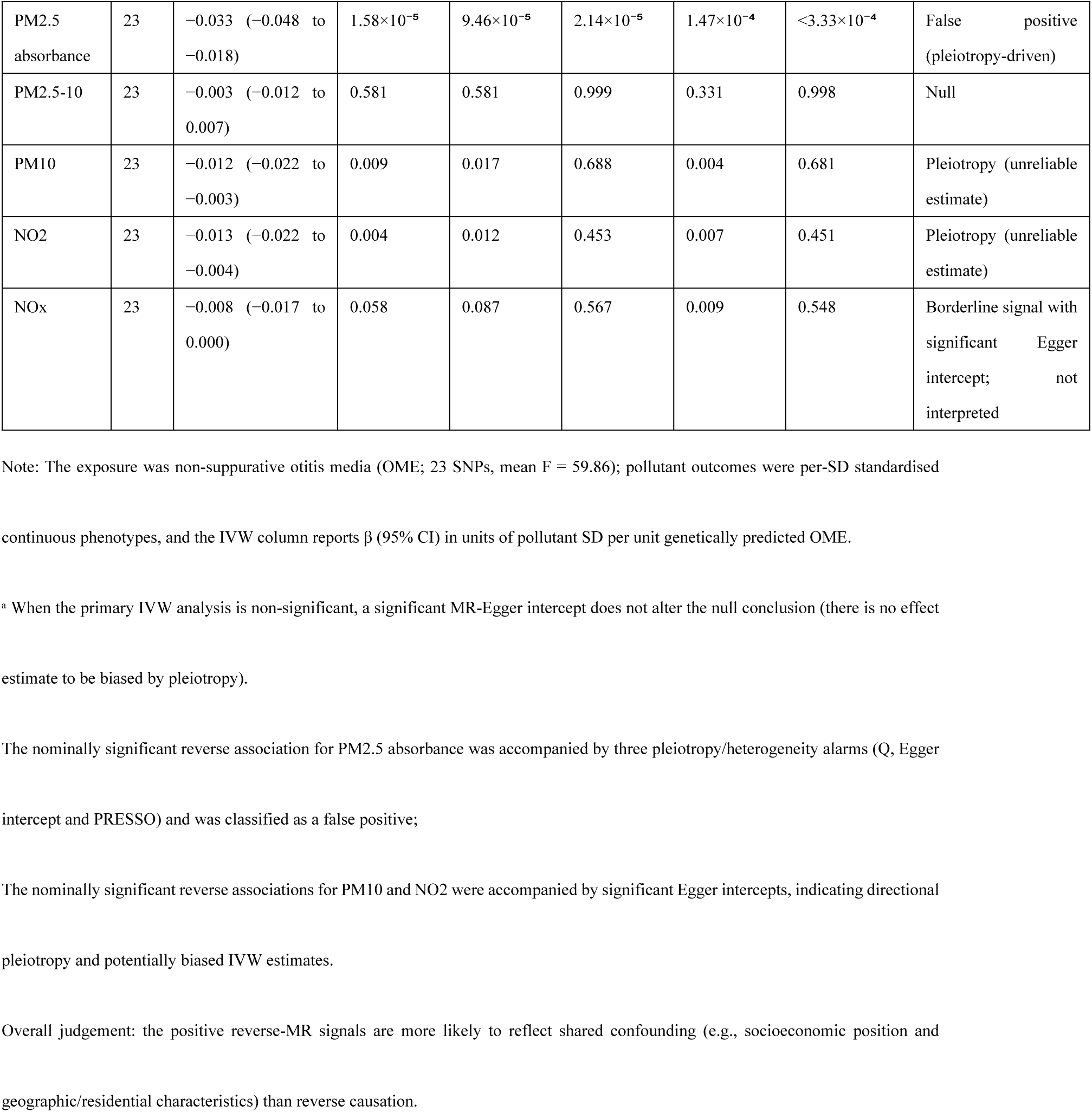
Reverse MR estimates for the association of non-suppurative otitis media with six air pollutants

| Outcome (pollutant) | SNP | IVW $\beta$ (95% CI, per SD) | P | FDR | Q (IVW) P | Egger intercept P | PRESSO global P | Overall classification |
| --- | --- | --- | --- | --- | --- | --- | --- | --- |
| PM2.5 | 23 | 0.006 (−0.003 to 0.015) | 0.226 | 0.271 | 0.829 | 0.027 | 0.833 | Null (non-significant IVW) |
| PM2.5 absorbance | 23 | −0.033 (−0.048 to −0.018) | $1.58 \times 10^{-5}$ | $9.46 \times 10^{-5}$ | $2.14 \times 10^{-5}$ | $1.47 \times 10^{-4}$ | $< 3.33 \times 10^{-4}$ | False positive (pleiotropy-driven) |
| PM2.5-10 | 23 | −0.003 (−0.012 to 0.007) | 0.581 | 0.581 | 0.999 | 0.331 | 0.998 | Null |
| PM10 | 23 | −0.012 (−0.022 to −0.003) | 0.009 | 0.017 | 0.688 | 0.004 | 0.681 | Pleiotropy (unreliable estimate) |
| NO2 | 23 | −0.013 (−0.022 to −0.004) | 0.004 | 0.012 | 0.453 | 0.007 | 0.451 | Pleiotropy (unreliable estimate) |
| NOx | 23 | −0.008 (−0.017 to 0.000) | 0.058 | 0.087 | 0.567 | 0.009 | 0.548 | Borderline signal with significant Egger intercept; not interpreted |

Note: The exposure was non-suppurative otitis media (OME; 23 SNPs, mean F = 59.86); pollutant outcomes were per-SD standardised
continuous phenotypes, and the IVW column reports $\beta$ (95% CI) in units of pollutant SD per unit genetically predicted OME.
<sup>a</sup> When the primary IVW analysis is non-significant, a significant MR-Egger intercept does not alter the null conclusion (there is no effect estimate to be biased by pleiotropy).
The nominally significant reverse association for PM2.5 absorbance was accompanied by three pleiotropy/heterogeneity alarms (Q, Egger intercept and PRESSO) and was classified as a false positive;
The nominally significant reverse associations for PM10 and NO2 were accompanied by significant Egger intercepts, indicating directional pleiotropy and potentially biased IVW estimates.
Overall judgement: the positive reverse-MR signals are more likely to reflect shared confounding (e.g., socioeconomic position and geographic/residential characteristics) than reverse causation.

### Exploratory multivariable sensitivity analyses

In the exploratory MVMR using the 88 SNPs with complete data, the conditional F-statistics for PM10 were below 10 in every model (6.54–8.50), and instrument strength on the covariate side was also insufficient (in the four-exposure model, conditional F was 9.17 for allergic rhinitis and 10.86 for asthma, below or near 10, with only chronic rhinosinusitis reaching 14.20), indicating weak-instrument problems on both the exposure and covariate sides. The MVMR-IVW estimates were unstable in direction across models (OR ≈ 0.90–2.31) and contradicted the pooled-set univariable estimate (OR = 0.64). Given conditional F < 10 and unstable effect directions, MVMR did not contribute to causal conclusions (complete estimates are provided in Table S4, conditional F distributions in Figure S5, and the models retained as exploratory analyses in Table 6 and Figure 5), and the null univariable MR estimates remain the primary basis for inference.

**Fig 5.**
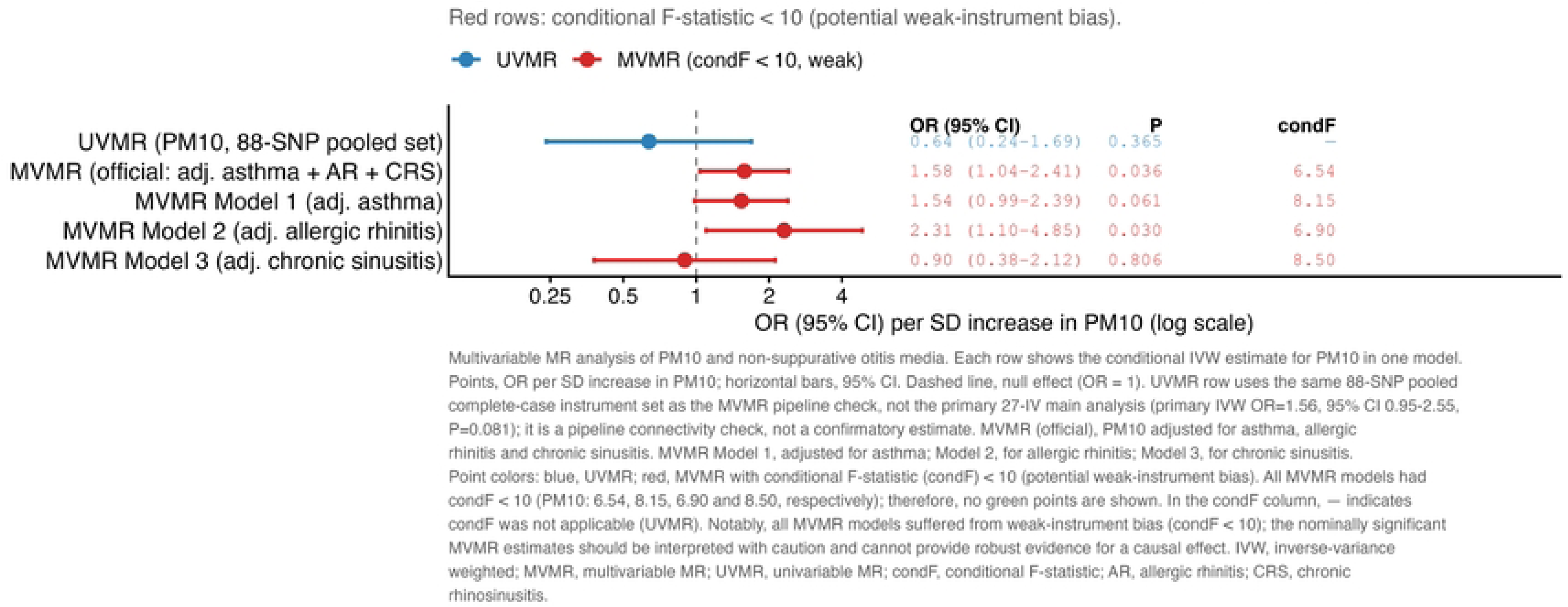
Conditional effect of PM10 in univariable and multivariable MR. Conditional IVW ORs (95% CIs) for PM10 are shown for the univariable 88-SNP pooled set (UVMR) and for multivariable models (MVMR) adjusted for asthma, allergic rhinitis and chronic rhinosinusitis (official model) and for each comorbidity separately (Models 1–3). All MVMR models had conditional F-statistics (condF) < 10, indicating weak-instrument bias; the nominally significant MVMR estimates are therefore not robust evidence. The UVMR pooled-set estimate (OR 0.64) is a pipeline connectivity check rather than the primary 27-IV analysis.

**Table 6.** Univariable and exploratory multivariable MR estimates for the association of PM10 with non-suppurative otitis media

| Model | Adjusted variables | SNP | PM10 conditional F | PM10 OR (95% CI) | P† | Covariate OR (95% CI) | Covariate conditional F |
| --- | --- | --- | --- | --- | --- | --- | --- |
| 88-SNP pooled-set UVMR (reference)<sup>a</sup> | None | 88 | — | 0.638 (0.241–1.689) | 0.365 | — | — |
| Four-exposure model | Asthma + rhinitis + rhinosinusitis | 88 | 6.54 | 1.579 (1.037–2.405) | 0.036† | Asthma 1.683 (1.554–1.822); rhinitis 1.010 (0.955–1.067); rhinosinusitis 0.883 (0.837–0.931) | 10.86; 9.17; 14.20 |
| Model 1 | Asthma | 88 | 8.15 | 1.537 (0.987–2.394) | 0.061 | Asthma 1.536 (1.469–1.607) | 81.05 |
| Model 2 | Allergic rhinitis | 88 | 6.90 | 2.308 (1.099–4.848) | 0.030† | Rhinitis 1.358 (1.274–1.449) | 31.47 |
| Model 3 | Chronic rhinosinusitis | 88 | 8.50 | 0.897 (0.380–2.121) | 0.806 | Rhinosinusitis 1.238 (1.144–1.340) | 47.37 |

Note: <sup>a</sup> The univariable UVMR reference was calculated from the pooled candidate SNP set used for MVMR (120 candidate SNPs; 115
retained after harmonisation):
only 88 SNPs had complete data for PM10 owing to its lower hit rate in the outcome (the four-exposure MVMR model used the same 88 SNPs), a different IV set from the primary analysis (27 SNPs after harmonisation; 30 extracted). The direction of this 88-SNP UVMR estimate (OR = 0.64) was opposite to that of the primary 27-SNP IVW estimate (OR = 1.555, 95% CI 0.947–2.553, P = 0.081), demonstrating that the pooled set contains weak instruments for PM10 and yields unstable results; the primary inference therefore always relies on the 27-SNP IVW analysis, and this row serves only as an internal MVMR reference.
Interpretation of MVMR results: †P values come from weak-instrument models with conditional F < 10 and are descriptive only; they cannot support causal interpretation. The Sanderson conditional F-statistics for PM10 were below 10 in all MVMR models (6.54–8.50), indicating weak-instrument bias; the direction of the PM10 effect was inconsistent across the four models (OR range 0.90–2.31) and contradicted the univariable UVMR estimate (OR = 0.64), and is judged unreliable.
Redistribution of covariate effects in MVMR: the estimates for the three covariates differed in direction and magnitude between MVMR and univariable analyses (e.g., chronic rhinosinusitis changed from a univariable OR = 1.28 to an MVMR OR = 0.88), indicating shared genetic structure among covariates whose effects were redistributed in MVMR; this further supports not using the MVMR results as causal evidence; the univariable MR results remain the primary basis for conclusions. This study does not claim an independent causal effect of PM10 on non-suppurative otitis media; causal inference for PM10 rests primarily on the (null) univariable MR results.
Abbreviations: MVMR, multivariable Mendelian randomization; UVMR, univariable Mendelian randomization.

## Discussion

Using a two-sample MR framework, this study systematically evaluated the potential causal associations of six air pollutants with non-suppurative otitis media (OME). The main findings are as follows. After FDR correction, none of the six pollutants was significantly causally associated with OME; only PM10 showed a nominal positive suggestion (OR = 1.555, P = 0.081), but this signal did not survive FDR correction, reversed in the univariable estimate from the 88-SNP pooled set (OR = 0.638, 95% CI 0.241–1.689) and was unstable in direction across MVMR models, indicating high sensitivity to instrument selection and model specification. Overall sensitivity analyses were robust: the Steiger test supported the pollutant → OME direction and leave-one-out analyses showed no single dominant SNP, with the sole exception of PM2.5 absorbance, which had only four instruments and significant heterogeneity and does not support directional conclusions. The nominally significant reverse-MR signals were all accompanied by significant Egger intercepts, and PM2.5 absorbance additionally showed significant heterogeneity and PRESSO tests, indicating horizontal pleiotropy and shared confounding rather than reverse causation. The MVMR analysis for PM10 had no causal-inference value owing to conditional F < 10 and unstable directions.

The predominantly null findings of this study contrast with previous observational epidemiological reports. Cohort and registry studies have linked prenatal or postnatal particulate matter and NO2 exposure to increased incidence, recurrence or hospitalisation of childhood otitis media^[4][7]^; systematic reviews have also reported positive associations between air pollution and childhood otitis media^[6]^, but the strength of association varies markedly across pollutants and studies, with some reporting only weak or non-robust associations^[8]^. Observational designs can rarely fully control for confounding by socioeconomic position, living environment, smoking, allergy and infection season, and can seldom disentangle the temporal order of exposure and onset^[30]^. Exploiting the quasi-natural experimental property of random allele allocation, this study offers genetic triangulation evidence for understanding these discrepancies^[31][32]^: observational studies provide real-world correlational clues, whereas MR relies on genetic instruments independent of postnatal confounding to set an upper bound of evidence for direct, lifetime, large-effect causal effects of pollutants on OME; the two designs have different bias sources and do not negate each other, and a null MR result with low power should not be directly interpreted as a correction of observational effect sizes^[33]^. In the absence of evidence for large-effect genetic causation, the positive associations reported observationally may partly arise from residual confounding or shared susceptibility to comorbidities such as allergy and asthma^[9][34][35]^, or from short-term triggering by pollutants during acute infection episodes^[7]^, rather than direct, lifelong, chronic causal effects on OME. This inference is consistent with the current MR evidence landscape: genetic causal effects of intrinsic immune factors such as BMI and allergic rhinitis on otitis media are relatively clear^[13][14]^, and MR studies of air pollution have likewise found no significant association for inner-ear disease (sudden sensorineural hearing loss)^[15]^, while genetic evidence for middle-ear OME remains absent. The mechanistic hypothesis that pollutants induce inflammatory oedema of the nasopharyngeal mucosa, impair Eustachian tube ventilation and drainage and cause negative middle-ear pressure and effusion is biologically plausible, and animal experiments offer some support^[5]^; however, MR uses genetic variants as proxies for lifetime-average exposure^[36]^, making it difficult to capture developmental susceptibility in infants and young children—the high-incidence window for OME^[4][37]^—and the short-term effects of high-concentration, seasonal acute exposure^[7][38]^, which are precisely the settings in which observational associations are most often reported. The null findings therefore do not rule out these mechanisms operating in specific exposure windows or at high doses; if genuine associations exist, they are more likely to be small-to-moderate effects within specific time windows than lifelong chronic large effects^[36][39]^. In addition, the uniformly null results for the three negative-control outcomes indicate only that no effect was detected at the available power and do not constitute evidence excluding broad non-specific pleiotropy^[40]^.

This study has several limitations. First, limited statistical power is the principal shortcoming: apart from PM2.5-10 and PM10, most exposures had only 4–7 instruments after harmonisation; PM2.5 absorbance (a black-carbon proxy) had only 4 SNPs after harmonisation with significant heterogeneity, and the MR-Egger intercept and MR-PRESSO tests have very low power with so few instruments, so its extremely wide confidence intervals do not support directional conclusions; larger samples and more source-specific black-carbon instruments are needed for re-evaluation. The threshold for PM2.5-10 and PM10 was relaxed to 5×10⁻⁶ because of insufficient genome-wide significant loci, which may introduce weaker instruments and potential pleiotropy; although all mean F values were >20, the F > 10 rule of thumb can be misleading^[41]^. We also did not systematically query external trait databases such as PhenoScanner for associations of every instrument with potential confounders, so instrument validity rests on the pleiotropy, directionality and control analyses rather than exhaustive SNP-level confounder screening. At 80% power, minimum detectable ORs ranged from 1.69 to 32.09 (Table 1). Second, the exposure GWAS came from UK Biobank in the UK and the outcome from FinnGen in Finland, which helps reduce bias related to sample overlap^[42]^, but both were European populations, and FinnGen derives from the genetically isolated Finnish population whose allele frequencies and linkage-disequilibrium structure differ subtly from those of Western European populations, limiting generalisation to non-European populations and regions with different environmental exposure backgrounds^[43]^. Third, the six pollutants are highly collinear in their sources and spatial distribution, making it difficult for univariable MR to isolate the effect of any single pollutant^[44]^; MVMR was in turn limited by weak conditional instruments and did not provide reliable independent-effect estimates. Fourth, pollutant exposure was estimated at residential addresses by a land-use regression model calibrated to 2010, entailing measurement error^[45]^, and the exclusion-restriction assumption of MR cannot be fully verified. Fifth, phenotype misclassification: H8_NONSUPPNAS is based on registry diagnostic codes and may mix heterogeneous non-suppurative phenotypes such as otitis media with effusion. We repeated the analyses for all six exposures using the three suppurative comparator phenotypes (acute, chronic and pooled), with consistent directions (all null), suggesting the main conclusion does not depend on a single phenotype definition; however, ICD codes could not be further disaggregated with summary data, and code-based phenotypes often have insufficient granularity and non-random missingness^[46]^; non-differential phenotype misclassification generally biases effect estimates towards the null, consistent with the conservative interpretation of the null findings. Together these factors argue for a conservative reading of the null results, and future life-course time-point-specific MR could test effects in developmental exposure windows^[39]^.

In summary, this study found no genetic evidence supporting large-effect causal associations of the six air pollutants with non-suppurative otitis media. Owing to limited statistical power, moderate effects of OR ≈ 1.2–1.5 cannot be excluded; the nominal PM10 signal is sensitive to instrument selection and does not constitute causal evidence, and its potential association particularly warrants further testing in larger samples with stronger, more specific instruments and multipollutant joint models. This study provides conservative, reproducible MR evidence for causal inference on air pollution and OME, suggesting that positive associations from observational studies should be interpreted cautiously in public-health communications, with due consideration of residual confounding, shared comorbidity and differences in exposure windows.

## Acknowledgments

The authors thank the UK Biobank, the MRC-IEU OpenGWAS consortium and FinnGen for making the GWAS summary data publicly available.

## Author contributions

Ling Yang conceived and designed the study, curated the data, performed formal analyses and wrote the original draft (Conceptualization, Data curation, Formal analysis, Writing–original draft); Yang Liu provided methodological guidance, supervised the study, reviewed and revised the manuscript and administered the project (Methodology, Supervision, Writing–review & editing, Project administration). All authors read and approved the final version of the manuscript.

## Competing interests

The authors declare that they have no competing interests.

## Data availability

This study used only publicly available GWAS summary statistics and did not access individual-level data. Air pollutant GWAS were obtained from the MRC-IEU OpenGWAS database (https://gwas.mrcieu.ac.uk/; GWAS IDs: ukb-b-10817, ukb-b-11312, ukb-b-12963, ukb-b-18469, ukb-b-9942, ukb-b-12417), and otitis media, comorbidity and control outcome data from FinnGen R13 (https://r13.finngen.fi/). All analyses were performed in R 4.5.2 using the TwoSampleMR (0.7.9) and MRPRESSO (1.0) packages; all analysis scripts, intermediate results and the STROBE-MR checklist are archived at Zenodo (https://doi.org/10.5281/zenodo.22892731) to ensure reproducibility.

## Ethics statement

As a secondary analysis of publicly available summary statistics without individual-level data, this study required no ethical approval or informed consent.

## Supporting information

**S1 Checklist.** STROBE-MR checklist completed with the page number and relevant text for each item.

## Notes

### Competing Interest Statement

The authors have declared no competing interest.

### Author Declarations

This study is a secondary analysis of publicly available genome-wide association study (GWAS) summary statistics. No individual-level data were accessed, and the study did not involve human participants, human specimens or tissue, vertebrate animals or cephalopods, vertebrate embryos or tissues, or field research. Therefore, no institutional review board (IRB) approval or ethical exemption was required. The original GWAS studies had obtained their own ethical approvals and informed consent. All data used are de-identified and publicly available.

## References

1. Rosenfeld RM, Shin JJ, Schwartz SR, et al. Clinical practice guideline: otitis media with effusion (update). Otolaryngol Head Neck Surg. 2016;154(1 Suppl):S1–S41. doi:10.1177/0194599815623467.

2. Chen R, Deng J, Sun Y, et al. Global, regional and national burdens of otitis media in children and adolescents from 1990 to 2021 and its predictions to 2040. Front Public Health. 2025;13:1552405. doi:10.3389/fpubh.2025.1552405.

3. Klimek L, Brough HA, Arasi S, et al. Otitis Media With Effusion (OME) and Eustachian Tube Dysfunction: The Role of Allergy and Immunity—An EAACI Position Paper. Allergy. 2025;80(9):2429–2441. doi:10.1111/all.16554.

4. Lu C, Li Q, Qiao Z, et al. Effects of pre-natal and post-natal exposures to air pollution on onset and recurrence of childhood otitis media. J Hazard Mater. 2023;459:132254. doi:10.1016/j.jhazmat.2023.132254.

5. Lee HM, Son YS, Kim HS, et al. Effects of particulate matter exposure on the eustachian tube and middle ear mucosa of rats. Clin Exp Otorhinolaryngol. 2023;16(3):225–235. doi:10.21053/ceo.2023.00227.

6. Bowatte G, Tham R, Perret JL, et al. Air pollution and otitis media in children: a systematic review of literature. Int J Environ Res Public Health. 2018;15(2):257. doi:10.3390/ijerph15020257.

7. Li C, Jiang X, Wei Y, et al. Air pollutants, seasonal influenza, and acute otitis media in children: a population-based analysis using 22-year hospitalization data. BMC Public Health. 2024;24(1):1581. doi:10.1186/s12889-024-18962-4.

8. MacIntyre EA, Karr CJ, Koehoorn M, et al. Residential air pollution and otitis media during the first two years of life. Epidemiology. 2011;22(1):81–89. doi:10.1097/EDE.0b013e3181fdb60f.

9. Wang YC, Yu YC, Chen SH, et al. Increased incidence of surgical intervention for otitis media with effusion among patients with type 2 inflammatory diseases. J Allergy Clin Immunol Pract. 2025;13(3):658–669. doi:10.1016/j.jaip.2024.12.014.

10. Allen EK, Chen WM, Weeks DE, et al. A genome-wide association study of chronic otitis media with effusion and recurrent otitis media identifies a novel susceptibility locus on chromosome 2. J Assoc Res Otolaryngol. 2013;14(6):791–800. doi:10.1007/s10162-013-0411-2.

11. Richmond RC, Davey Smith G. Mendelian randomization: Concepts and scope. Cold Spring Harb Perspect Med. 2022;12(1):a040501. doi:10.1101/cshperspect.a040501.

12. Burgess S, Butterworth A, Thompson SG. Mendelian randomization analysis with multiple genetic variants using summarized data. Genet Epidemiol. 2013;37(7):658–665. doi:10.1002/gepi.21758.

13. Cao J, Liu W, Yang Z, et al. Causal relationship between body mass index and risk of otitis media with effusion in children: a Mendelian randomization study. Indian J Otolaryngol Head Neck Surg. 2024;76(1):1410–1418. doi:10.1007/s12070-023-04161-x.

14. Wang F, Yu C, Liu R. Causal relationship between allergic rhinitis and otitis media: a Mendelian randomization study. Medicine (Baltimore). 2024;103(39):e39671. doi:10.1097/MD.0000000000039671.

15. Luo X, Zuo W, Ren Q, et al. Correlation of air pollution and risk of sudden sensorineural hearing loss: a Mendelian randomization study. Sci Rep. 2025;15(1):28921. doi:10.1038/s41598-025-92952-3.

16. Yang S, Tong T, Wang H, et al. Causal relationship between air pollution and infections: a two-sample Mendelian randomization study. Front Public Health. 2024;12:1409640. doi:10.3389/fpubh.2024.1409640.

17. Chen F, Zhang J, Xie J, et al. Causal relationships between air pollutants and upper respiratory tract infections: a two-sample Mendelian randomization study. J Chin Med Assoc. 2025;88(6):481–485. doi:10.1097/JCMA.0000000000001242.

18. Yu Y, Yuan R, Zhang J, et al. Systems perspectives on pediatric otitis media: environmental exposures, genetic susceptibility, and biomarker-guided interventions. Front Pediatr. 2026;14:1758098. doi:10.3389/fped.2026.1758098.

19. Skrivankova VW, Richmond RC, Woolf BAR, et al. Strengthening the reporting of observational studies in epidemiology using Mendelian randomization: the STROBE-MR statement. JAMA. 2021;326(16):1614–1621. doi:10.1001/jama.2021.18236.

20. Hemani G, Zheng J, Elsworth B, et al. The MR-base platform supports systematic causal inference across the human phenome. Elife. 2018;7:e34408. doi:10.7554/eLife.34408.

21. Sudlow C, Gallacher J, Allen N, et al. UK Biobank: An open access resource for identifying the causes of a wide range of complex diseases of middle and old age. PLoS Med. 2015;12(3):e1001779. doi:10.1371/journal.pmed.1001779.

22. Kurki MI, Karjalainen J, Palta P, et al. FinnGen provides genetic insights from a well-phenotyped isolated population. Nature. 2023;613(7944):508–518. doi:10.1038/s41586-022-05473-8.

23. Eeftens M, Beelen R, De Hoogh K, et al. Development of land use regression models for PM(2.5), PM(2.5) absorbance, PM(10) and PM(coarse) in 20 European study areas; results of the ESCAPE project. Environ Sci Technol. 2012;46(20):11195–11205. doi:10.1021/es301948k.

24. de Hoogh K, Gulliver J, van Donkelaar A, et al. Development of West-European PM2.5 and NO2 land use regression models incorporating satellite-derived and chemical transport modelling data. Environ Res. 2016;151:1–10. doi:10.1016/j.envres.2016.07.005.

25. Bowden J, Davey Smith G, Haycock PC, et al. Consistent estimation in Mendelian randomization with some invalid instruments using a weighted median estimator. Genet Epidemiol. 2016;40(4):304–314. doi:10.1002/gepi.21965.

26. Bowden J, Davey Smith G, Burgess S. Mendelian randomization with invalid instruments: effect estimation and bias detection through Egger regression. Int J Epidemiol. 2015;44(2):512–525. doi:10.1093/ije/dyv080.

27. Verbanck M, Chen CY, Neale B, et al. Detection of widespread horizontal pleiotropy in causal relationships inferred from Mendelian randomization between complex traits and diseases. Nat Genet. 2018;50(5):693–698. doi:10.1038/s41588-018-0099-7.

28. Sanderson E, Windmeijer F. A weak instrument F-test in linear IV models with multiple endogenous variables. J Econom. 2016;190(2):212–221. doi:10.1016/j.jeconom.2015.06.004.

29. Sanderson E, Davey Smith G, Windmeijer F, et al. An examination of multivariable Mendelian randomization in the single-sample and two-sample summary data settings. Int J Epidemiol. 2019;48(3):713–727. doi:10.1093/ije/dyy262.

30. Davies NM, Holmes MV, Davey Smith G. Reading Mendelian randomisation studies: a guide, glossary, and checklist for clinicians. BMJ. 2018;362:k601. doi:10.1136/bmj.k601.

31. Lawlor DA, Tilling K, Davey Smith G. Triangulation in aetiological epidemiology. Int J Epidemiol. 2016;45(6):1866–1886. doi:10.1093/ije/dyw314.

32. Munafò MR, Davey Smith G. Robust research needs many lines of evidence. Nature. 2018;553(7689):399–401. doi:10.1038/d41586-018-01023-3.

33. Burgess S, Davey Smith G, Davies NM, et al. Guidelines for performing Mendelian randomization investigations: update for summer 2023. Wellcome Open Res. 2023;4:186. doi:10.12688/wellcomeopenres.15555.3.

34. Skoner AR, Skoner KR, Skoner DP. Allergic rhinitis, histamine, and otitis media. Allergy Asthma Proc. 2009;30(5):470–481. doi:10.2500/aap.2009.30.3272.

35. Kreiner-Møller E, Chawes BLK, Caye-Thomasen P, et al. Allergic rhinitis is associated with otitis media with effusion: A birth cohort study. Clin Exp Allergy. 2012;42(11):1615–1620. doi:10.1111/j.1365-2222.2012.04038.x.

36. Labrecque JA, Swanson SA. Interpretation and potential biases of Mendelian randomization estimates with time-varying exposures. Am J Epidemiol. 2019;188(1):231–238. doi:10.1093/aje/kwy204.

37. Oh J, Lee S, Kim MH, et al. The impact of PM2.5 on acute otitis media in children (aged 0-3): A time series study. Environ Int. 2020;145:106133. doi:10.1016/j.envint.2020.106133.

38. Park M, Han J, Park J, et al. Particular matter influences the incidence of acute otitis media in children. Sci Rep. 2021;11(1):19730. doi:10.1038/s41598-021-99247-3.

39. Sanderson E, Richardson TG, Morris TT, et al. Estimation of causal effects of a time-varying exposure at multiple time points through multivariable Mendelian randomization. PLoS Genet. 2022;18(7):e1010290. doi:10.1371/journal.pgen.1010290.

40. Lipsitch M, Tchetgen Tchetgen E, Cohen T. Negative controls: a tool for detecting confounding and bias in observational studies. Epidemiology. 2010;21(3):383–388. doi:10.1097/EDE.0b013e3181d61eeb.

41. Burgess S, Thompson SG; CRP CHD Genetics Collaboration. Avoiding bias from weak instruments in Mendelian randomization studies. Int J Epidemiol. 2011;40(3):755–764. doi:10.1093/ije/dyr036.

42. Burgess S, Davies NM, Thompson SG. Bias due to participant overlap in two-sample Mendelian randomization. Genet Epidemiol. 2016;40(7):597–608. doi:10.1002/gepi.21998.

43. Wojcik GL, Graff M, Nishimura KK, et al. Genetic analyses of diverse populations improves discovery for complex traits. Nature. 2019;570(7762):514–518. doi:10.1038/s41586-019-1310-4.

44. Dominici F, Peng RD, Barr CD, et al. Protecting human health from air pollution: Shifting from a single-pollutant to a multipollutant approach. Epidemiology. 2010;21(2):187–194. doi:10.1097/EDE.0b013e3181cc86e8.

45. Basagaña X, Aguilera I, Rivera M, et al. Measurement error in epidemiologic studies of air pollution based on land-use regression models. Am J Epidemiol. 2013;178(8):1342–1346. doi:10.1093/aje/kwt127.

46. Bush WS, Oetjens MT, Crawford DC. Unravelling the human genome-phenome relationship using phenome-wide association studies. Nat Rev Genet. 2016;17(3):129–145. doi:10.1038/nrg.2015.36.

